# A retrospective cohort study investigating synergism of air pollution and corticosteroid exposure in promoting cardiovascular and thromboembolic events in older adults

**DOI:** 10.1101/2022.12.15.22283489

**Authors:** Kevin P. Josey, Rachel C. Nethery, Aayush Visaria, Benjamin Bates, Poonam Gandhi, Ashwaghosha Parthasarathi, Melanie Rua, David Robinson, Soko Setoguchi

**Author notes:** Rutgers University Institute for Health, Healthcare Policy, and Aging Research 112 Paterson Street, Room 402 New Brunswick, NJ 08901, USA.

## Abstract

**Objective:** To evaluate the synergistic effects created by fine particulate matter (PM_2.5_) and corticosteroid use on hospitalization and mortality in older adults at high-risk for cardiovascular thromboembolic events (CTEs).

**Design and Setting:** A retrospective cohort study using a US nationwide administrative healthcare claims database.

**Participants:** A 50% random sample of participants with high-risk conditions for CTE from the 2008-2016 Medicare Fee-for-Service population.

**Exposures:** Corticosteroid therapy and seasonal-average PM_2.5_.

**Main Outcome Measures:** Incidences of myocardial infarction or acute coronary syndrome, ischemic stroke or transient ischemic attack, heart failure, venous thromboembolism, atrial fibrillation, and all-cause mortality. We assessed additive interactions between PM_2.5_ and corticosteroids using estimates of the relative excess risk due to interaction (RERI) obtained using marginal structural models for causal inference.

**Results:** Among the 1,936,786 individuals in the high CTE risk cohort (mean age 76.8, 40.0% male, 87.4% White), the mean PM_2.5_ exposure level was 8.3 ± 2.4 μg/m^3^ and 37.7% had at least one prescription for a systemic corticosteroid during follow-up. For all outcomes, we observed increases in risk associated with corticosteroid use and with increasing PM_2.5_ exposure. PM_2.5_ demonstrated a non-linear relationship with some outcomes. We also observed evidence of an interaction existing between corticosteroid use and PM_2.5_ for some CTEs. For an increase in PM_2.5_ from 8 μg/m^3^ to 12 μg/m^3^ (a policy-relevant change), the RERI of corticosteroid use and PM_2.5_ was significant for heart failure (15.6%, 95% CI: 4.0%-27.3%). Increasing PM_2.5_ from 5 μg/m^3^ to 10 μg/m^3^ yielded significant RERIs for incidences of heart failure (32.4; 95% CI: 14.9%-49.9%) and myocardial infarction/acute coronary syndromes (29.8%; 95% CI: 5.5%-54.0%).

**Conclusion:** PM_2.5_ and systemic corticosteroid use were independently associated with increases in CTE hospitalizations. We also found evidence of significant additive interactions between the two exposures for heart failure and myocardial infarction/acute coronary syndromes suggesting synergy between these two exposures.

**Strengths and Limitations of this Study:**

- We conduct analyses using robust causal inference and machine learning techniques and incorporate a large set of individual-level factors that are typically absent in environmental health analyses with large claims data sets.
- We present statistics that evaluate the synergy between fine particulate matter and corticosteroid therapy on the additive scale, which is more relevant for assessing excess risks and informing policy.
- Patient medical history prior to receiving Medicare benefits is unknowable within a Fee-for-Service claims database, which may lead to exclusion of some high-risk individuals from the cohort.
- We censor participants after the earlier of the end of first corticosteroid regimen or the first incidence of the outcome of interest, which makes the analyses statistically tractable but sacrifices some information in the data.

## Introduction

Climate change is ‘the single biggest health threat facing humanity,’^1^ and is expected to have a growing impact on human health through multiple pathways, including more frequent extreme weather events and worsening ambient air pollution.^2^ Air pollution is currently among the top five modifiable contributors to death and disease globally.^3^ The impacts of air pollution, specifically fine particulate matter (PM_2.5_), on the cardiovascular system are well-established. PM_2.5_ exposure has been linked to increased risk of stroke, myocardial infarction, heart failure, venous thromboembolism, and other cardiovascular events.^4–11^ More than half of deaths attributable to air pollutants are due to cardiovascular thromboembolic (CTE) events.^12^ Epidemiological assessments in this area are also supported by cellular/toxicological experiments and by controlled animal/human studies, which both demonstrate the mechanisms by which PM_2.5_ may trigger acute events as well as prompt the chronic development of cardiovascular diseases.^13^

One of the most vulnerable populations, older adults, are at elevated risk for mortality and morbidity from PM_2.5_, particularly those with accessory comorbidities such as respiratory and cardiovascular diseases.^14^ Older adults are also at increased risk for CTE from certain medications taken to treat or prevent comorbidities.^15,16^ For example, systemic corticosteroids used for asthma/COPD exacerbations and to treat autoimmune diseases have direct vasoconstriction effects that inhibit fibrinolytic activity of the blood, leading to clinically-recognized thrombogenicity.^17^ Further, systemic corticosteroids can cause sodium and fluid retention issues, leading to hypertension or heart failure exacerbations.^18–20^

Although the independent effects of air pollution and corticosteroids on CTE are well-known, no prior study has assessed the risk of both exposures simultaneously on CTE. Thus, it is unknown whether there is a synergy between these factors. Leveraging rich healthcare utilization data on a large cohort of Medicare beneficiaries with comorbidities linked to residential PM_2.5_ concentrations, we examined whether simultaneously experiencing elevated PM_2.5_ concentrations and being exposed to corticosteroid therapies leads to an increased risk of CTE that is greater than the combination of these two effects independently. To our knowledge this is the first study to examine interactions between a drug and an air pollution exposure. We therefore provide a causal analytic framework that enables robust investigation of the contributing factors that explain individual-specific vulnerabilities to air pollution through the evaluation of additive interactions in survival models. Additionally, our analyses adjust for a large set of individual-level potential confounders that are typically unmeasured in environmental health analyses with large claims datasets, which lends added credibility to our findings.

## Methods

### Study Population and Cohort Definition

The cohort used in this study has been previously described.^14^ Briefly, we used data from a 50% random sample of the 2008-2016 Medicare Part D-eligible Fee-for-Service beneficiary population and formed a cohort of individuals with conditions known to increase the risk of CTE. These high-risk conditions included pre-existing cardiovascular diseases, prior venous thromboembolism, total joint arthroplasty, and cancer. Any beneficiary who had an inpatient diagnosis/procedure at any position (primary or otherwise) for one or more of the above causes during a one-year baseline period from their date of enrollment into the Medicare Fee-for-Service system was entered into the cohort at the end of the baseline period. This definition for high CTE risk has been shown to be highly predictive of future CTE events (see Supplemental Table S1 for specific International Classification of Diseases (ICD-9/10) diagnosis codes used to define each high-risk condition). ^21^

### Outcomes

We followed all participants until they developed one of the outcomes of interest, or until they experienced a censoring event – whichever occurred first. Outcomes of interest included hospitalization for: 1) myocardial infarction or acute coronary syndrome (MI/ACS); 2) ischemic stroke or transient ischemic attack (Stroke/TIA); 3) heart failure (HF); 4) atrial fibrillation (Afib); 5) venous thromboembolism (VTE); or 6) death from any cause (see Supplemental Table S2 for ICD-9/10 diagnosis and procedure codes identifying the outcomes). Non-administrative censoring events included death (when death was not the outcome under study), loss of eligibility for Medicare Part D, and the participant’s moving to a ZIP-code without available PM_2.5_ exposure data. We also censored participants after the discontinuation of their first corticosteroid therapy, with a 30-day grace period.

### PM_2.5_ Exposures

Seasonal-average PM_2.5_ concentrations were derived from spatially and temporally aggregated predictions from a well-validated, high-resolution PM_2.5_ model.^22^ This model predicts PM_2.5_ concentrations at 1-km square grids across the US and consists of an ensemble of neural net and machine learning sub-models trained on integrated high-resolution satellite, land use, emissions, ground monitoring, and weather data. Daily gridded estimates were aggregated and linked to participants by residential ZIP-code, and then averaged within seasons. Figure 1 demonstrates the significant between-season variation in PM_2.5_ patterns in the US, which motivated our choice to examine seasonal-average PM_2.5_ exposures rather than more traditional yearly-average exposures as shorter-term increases in exposure may have harmful effects in this vulnerable population. We are particularly focused on contrasting outcomes under PM_2.5_ exposures of 12 μg/m^3^ vs. 8 μg/m^3^, which are policy-relevant thresholds currently in review by the US Environmental Protection Agency (EPA; 12 μg/m^3^ being the current US limit for annual average PM_2.5_).^23^ We also compare outcomes for the contrast between PM_2.5_ levels of 10 μg/m^3^ vs. 5 μg/m^3^ in a secondary analysis, informed by the WHO’s updated guidelines recommending an annual average limit of 5 μg/m^3^ (recently reduced from 10 μg/m^3^).^24^

**Figure 1.**
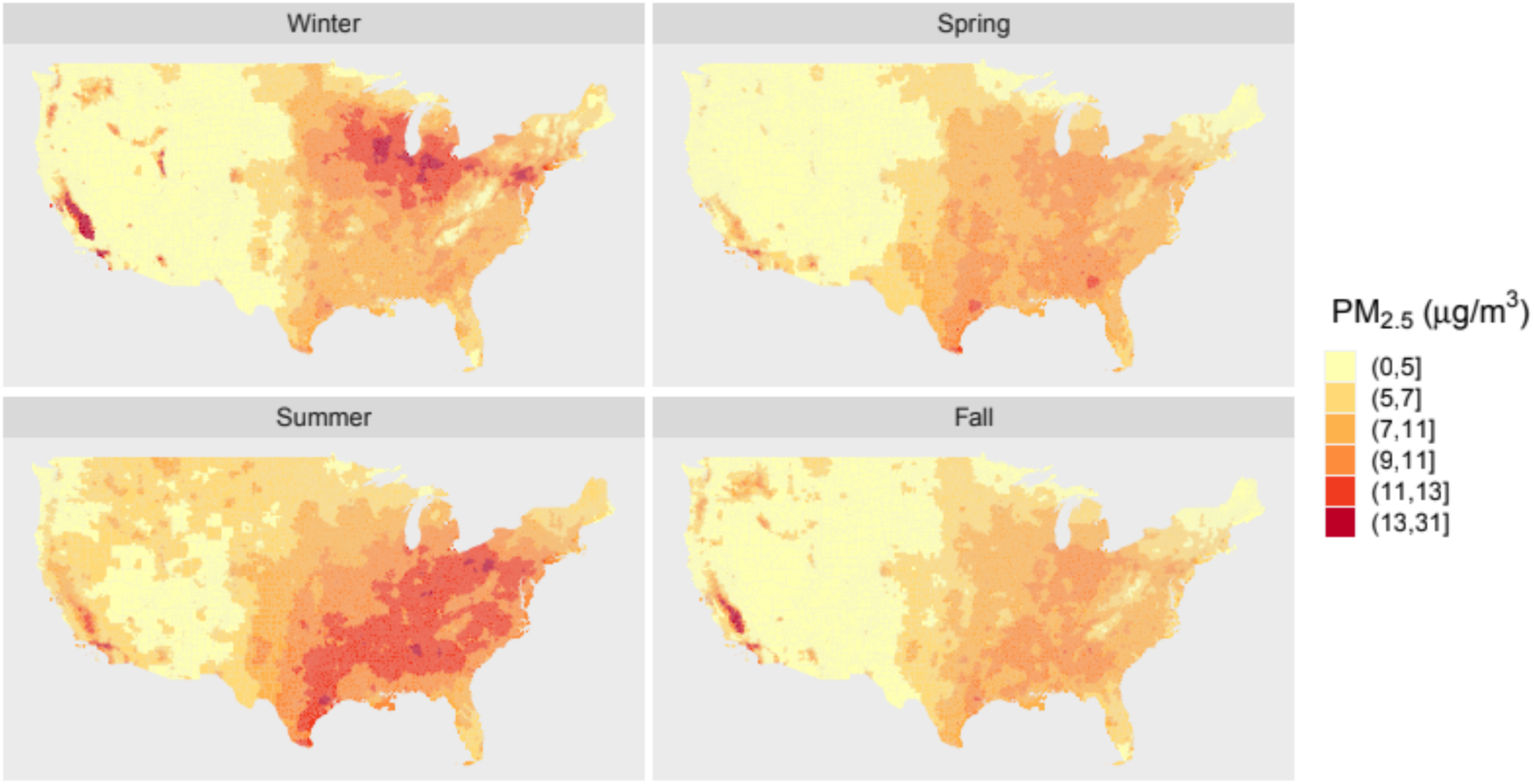
Season-specific average PM_2.5_ measurements for every ZIP-code across the United States over the period 2008-2016. Note that the PM_2.5_ measurements are most severe in the Southern states during the Summer and in the upper-Midwest states during Winter.

### Corticosteroid Exposures

We used Medicare Part D drug dispensing data to identify systemic corticosteroid exposure. Systemic corticosteroids of interest included Cortisone, Hydrocortisone, Prednisone, Prednisolone, Methylprednisolone, Triamcinolone, Dexamethasone, and Betamethasone. Initiation and duration of each corticosteroid were estimated based on the dispensing date, dispensing dose, and days’ supply of the participants’ prescriptions. Because allowing for continuous follow-up was computationally infeasible, corticosteroid therapy status was updated quarterly until one of the study endpoints was achieved for each participant.

To ensure that individuals’ quarterly follow-up times aligned with key dates of corticosteroid usage, we constructed unique drug exposure panels for each cohort member. The anchor point of the drug exposure panels is the date of initiation of corticosteroid therapy during follow-up for users. In other words, for a participant who uses corticosteroids at some point during follow-up, the first day of their corticosteroid therapy always coincides with the first day of a quarter. We then constructed individual-specific panels spanning quarterly intervals extending backward in time to the participant’s index date and forward in time to the participant’s end date (see Supplemental Figure S1 for example). For individuals who never use corticosteroids during follow-up, quarter start times coincide with changes in season.

### Covariates

We identified individual-level sociodemographic characteristics, comorbidities and health services utilization information derived from Medicare enrollment files and inpatient, outpatient, and drug dispensing data from files pertaining to Medicare Parts A, B, and D, respectively. Using Medicare enrollment files, we extracted the following individual-level baseline variables: age, sex, race/ethnicity, and Medicaid eligibility (a proxy for low-income status). Various pre-enrollment measurements were assessed based on diagnosis codes for inpatient and outpatient visits during each participant’s baseline period (see Tables S3 for complete list of comorbidities). We also derived metrics of health services utilization during the baseline period, including the number of hospitalizations, number of emergency department visits, number of outpatient visits, and number of generic medications dispensed. We consider this collection of variables as time-invariant and treat them as potential confounders between corticosteroid use and CTE.

Additional temporal and neighborhood-level features were also identified to enable further confounding adjustment, for both PM_2.5_-CTE and corticosteroid-CTE associations. These variables included season, year, region, and PM_2.5_ from the prior four seasons as well as area-based measures of population density, proportion of residents living below the federal poverty line, proportion of housing units that are owner-occupied, median home value, median household income, proportion of residents identifying as Hispanic, proportion of residents identifying as Black, and proportion of residents 25+ with at least a college degree that were linked to Medicare by ZIP-code of beneficiaries’ residence. These demographic and socioeconomic features were considered as time-varying covariates updated yearly. We also accounted for ZIP-code changes that occur during follow-up and updated participants’ PM_2.5_ exposures and neighborhood features accordingly.

### Statistical Analysis

We first described summary measures of the individual-and neighborhood-level characteristics and calculated the number of person-years at risk, number of events, and event rates per 1,000 person years for each of the six outcomes examined, both overall and stratified by corticosteroid status. We then fitted history-adjusted marginal structural Cox proportional hazard models, facilitated by estimated inverse probability weights (IPWs), to investigate both the independent and synergistic effects of PM_2.5_ and corticosteroid use on the CTE outcomes.^25,26^ Separate models were used to estimate the IPWs for each of the outcomes considered, and separate weighted Cox models were fit over the age-time scale.^27^ We included penalized spline components in the weighted Cox models to account for potential nonlinear effects of PM_2.5_, in addition to a main effect for corticosteroid use and an interaction between corticosteroid use and PM_2.5_ (the penalized spline representation). Given that our cohort comprises individuals with diverse diseases each potentially affecting thrombosis risk, we employed stratification in the Cox models based on disease indication. Particularly, we allowed for disease-specific baseline hazards for autoimmune diseases and COPD/asthma (see Table S4 for ICD-9/10 codes), as these two categories of diseases exhibit unique pathways for cardiovascular and thromboembolic events independent of corticosteroid therapy.^28–33^

The final IPW for a given participant and follow-up period was constructed as the product of three distinct IPWs accounting for different potential sources of bias: an inverse probability of treatment weight for each of the PM_2.5_ and corticosteroid exposures, to adjust for confounding, and an inverse probability of censoring weight to account for informative censoring. The ZIP-code and season-specific IPWs for PM_2.5_ were constructed by taking the inverse of estimated generalized propensity scores modeled using gradient boosting regression. The individual-level IPWs for the quarterly corticosteroid use indicators were obtained by inverting propensity scores estimated using gradient boosting classification. Additionally, over the same corticosteroid use quarters, we modeled the probability of censoring with gradient boosting classification to produce inverse probability of censoring weights. The three IPWs were stabilized by the marginal probabilities of treatment/censoring. Extreme weights were truncated at the 1^st^ and 99^th^ percentiles of the final IPW distribution (see Supplement for additional details on the construction of the IPWs).

We report the hazard ratio estimates and 95% confidence intervals associating PM_2.5_ with the five CTE outcomes and all-cause mortality (comparing average hazards evaluated at PM_2.5_ levels of 12 vs. 8 μg/m^3^ and 10 vs. 5 μg/m^3^) with corticosteroid status held fixed (both on and off treatment). We also provide hazard ratios associating corticosteroid use with each outcome, with PM_2.5_ held fixed at 8 μg/m^3^. We assessed synergy between PM_2.5_ and corticosteroids by calculating the relative excess risk due to interaction (RERI) – a measure of interaction on the additive scale that can be interpreted as the relative increase in risk due to the combined effect of the two exposures versus the individual effects of the two exposures summed together (presented as a percentage).^34–38^ Cluster m-out-of-n bootstrap samples of the Cox model parameters were used to compute standard errors for the RERI which account for the correlation between units in the same ZIP-code while maintaining adequate computational efficiency.^39^ Additional details for estimating the RERI are provided in the Supplement.

This study was approved by the Institutional Review Board of Rutgers University. All analyses were conducted using R version 4.2.0. Data cleaning was performed using SAS version 9.4.

### Patient and public involvement

Patients and/or the public were not involved in the design, conduct, reporting, or dissemination of this research.

## Results

The cohort included 1,936,786 beneficiaries with a total of 4,629,432 person-years of follow-up. Average age at index date was 76.8 years, with 60.0% of cohort members female, 15.9% Medicaid eligible, 87.4% White, and 8.2% Black (Table 1). The average participant follow-up time was 2.4 ± 2.3 years, although this figure along with the total person-years of follow-up fluctuates depending on the outcome being evaluated (Supplemental Table S5). Among the Medicare beneficiaries evaluated, 37.7% had at least one prescription for corticosteroid therapy during follow-up. Participants who received corticosteroid therapy were slightly younger than those who never received corticosteroid therapy (75.7 versus 77.4 years old), were more likely to be White (89.8% versus 85.9%) and were less likely to be Medicaid eligible (14.0% versus 17.0%). Table 1 and Supplemental Table S3 shows summary statistics of several comorbidities included into the IPW models, stratified by disease indications listed in Table S4. Table 2 contains data on demographics and season-specific PM_2.5_ measurements over 329,544 ZIP-code years from 35,695 unique ZIP-codes. The average PM_2.5_ level was 8.3 ± 2.4 μg/m^3^, average population density was 1,425 people per square mile, and the overall poverty rate was 10.3%.

**Table 1.** Characteristics of high-risk Medicare beneficiaries (total N=1,936,786). Beneficiaries are further divided into strata of those who received at least one dispensing of systemic corticosteroid during follow-up and those who never received corticosteroids during follow-up. We report counts (%) for categorical variables and mean ± standard deviation for continuous variables.

| Variable | Level | All Participants<br>(N = 1,936,786) | Participants with<br>Autoimmune Diseases<br>(N = 212,697) | Participants with COPD or<br>Asthma (N = 510,941) |
| --- | --- | --- | --- | --- |
| Age at Index | | 76.8 ± 8.0 | 75.9 ± 7.3 | 76.5 ± 7.7 |
| Male |  | 774,852 (40.0) | 88,897 (41.8) | 204,382 (40.0) |
| Race |  |  |  |  |
|  | White | 1,692,791 (87.4) | 183,623 (86.3) | 449,203 (87.9) |
|  | Black | 159,001 (8.2) | 20,555 (9.7) | 41,194 (8.1) |
|  | Hispanic | 35,508 (1.8) | 3,110 (1.5) | 9,337 (1.8) |
|  | Asian | 24,152 (1.3) | 2,582 (1.2) | 5,293 (1.0) |
|  | North American<br>Native | 6,943 (0.4) | 649 (0.3) | 2,026 (0.4) |
|  | Other | 18,391 (1.0) | 2,178 (1.0) | 3,888 (0.8) |
| Medicaid Eligibility |  | 307,985 (15.9) | 29,050 (13.6) | 103,352 (20.2) |
| Chronic Conditions |  |  |  |  |
|  | Total Hip Arthroplasty | 231,114 (11.9) | 30,939 (14.6) | 54,452 (10.7) |
|  | Total Knee<br>Arthroplasty | 473,206 (24.4) | 66,549 (31.3) | 104,298 (20.4) |
|  | Acute Coronary<br>Syndrome | 74,783 (3.9) | 7,263 (3.4) | 21,984 (4.3) |
|  | Cancer | 291,014 (15.0) | 25,508 (12.0) | 76,843 (15.0) |
|  | Atrial Fibrillation | 595,876 (30.8) | 64,386 (30.3) | 177,324 (34.7) |
|  | Hemorrhagic Stroke | 184,442 (9.5) | 16,084 (7.6) | 45,901 (9.0) |
|  | Heart Failure | 291,993 (15.1) | 37,048 (17.4) | 111,890 (21.9) |
|  | Ischemic Stroke | 175,201 (9.1) | 15,453 (7.3) | 44,106 (8.6) |
|  | Myocardial Infarction | 127,147 (6.5) | 11,516 (5.4) | 38,020 (7.4) |
|  | Peripheral Vascular Disease | 196,055 (10.1) | 20,666 (9.7) | 68,855 (13.5) |
|  | Transient Ischemic Attack | 56,930 (2.9) | 5,073 (2.4) | 13,806 (2.7) |
|  | Venous Thromboembolism | 95,081 (4.9) | 11,035 (5.2) | 28,077 (5.5) |
|  | Cerebrovascular Accident | 184,442 (9.5) | 16,084 (7.6) | 45,901 (9.0) |
|  | Carotid Stenosis | 828 (0.0) | 100 (0.1) | 304 (0.1) |
| Hospitalization and Medication History |  |  |  |  |
|  | Prior Inpatient Hospitalizations | 1.7 ± 1.3 | 1.8 ± 1.4 | 2.0 ± 1.7 |
|  | Prior ER Visits | 0.8 ± 1.2 | 0.8 ± 1.3 | 1.1 ± 1.5 |
|  | Prior Outpatient Visits | 8.3 ± 9.2 | 8.9 ± 9.5 | 9.0 ± 9.5 |
|  | Prior Medications Dispensed | 12.1 ± 7.1 | 14.0 ± 7.4 | 14.8 ± 7.8 |

**Table 2.** Neighborhood-level characteristics averaged over 329,727 ZIP-code years across 35,695 unique ZIP-codes.

| Variable | Level | Mean ± SD |
| --- | --- | --- |
| PM<sub>2.5</sub> ($\mu\text{g}/\text{m}^3$) | | |
| | Yearly Average | 8.3 ± 2.4 |
| | Winter | 8.7 ± 3.4 |
| | Spring | 7.8 ± 2.4 |
| | Summer | 9.5 ± 2.9 |
| | Fall | 7.5 ± 2.4 |
| Population Density (per mile<sup>2</sup>) | | 1,425.0 ± 4,978.5 |
| Median House Value (\$) | | 180,117.4 ± 154,630.3 |
| Median Household Income (\$) | | 52,276.7 ± 22,549.9 |
| % Poverty | | 10.3 ± 10.7 |
| % Owner Occupied Housing | | 72.3 ± 17.4 |
| % Hispanic | | 9.7 ± 17.1 |
| % Black | | 8.5 ± 16.5 |
| % White | | 82.9 ± 20.8 |
| % less than High School | | 23.5 ± 16.8 |

During an average follow-up of 2.4 years per person, we observed a total of 244,451 hospitalization for HF, 118,754 hospitalizations for Afib, 101,611 hospitalizations for Stroke/TIA, 93,191 hospitalizations for MI/ACS, 41,635 hospitalizations for VTE, and 491,445 deaths. The incidence rates per 1,000 person-years were 57.1 for HF, 27.0 for Afib, 22.8 Stroke/TIA, 20.8 for MI/ACS, 9.1 for VTE, and 106.2 for death (Supplemental Table Table S5).

Corticosteroid use was associated with higher risks of CTE and death, with significant associations for all six outcomes examined. Holding PM_2.5_ fixed at 8 μg/m^3^, the hazard ratios (95% CI) for corticosteroid use were 2.04 (1.94, 2.14) for MI/ACS, 1.51 (1.42, 1.61) for Stroke/TIA, 2.18 (2.11, 2.25) for HF, 3.39 (3.19, 3.61) for VTE, 2.25 (2.11, 2.40) for Afib, and 2.64 (2.57, 2.72) for Death.

Seasonal-average PM_2.5_ exposure was also significantly associated with an increased risk of each of the six CTE and mortality outcomes. Increasing the PM_2.5_ concentration from 8 μg/m^3^ to 12 μg/m^3^, in the absence of corticosteroid therapy, resulted in hazard ratios (95% CI) of 1.244 (1.226, 1.263) for MI/ACS, 1.252 (1.234, 1.271) for Stroke/TIA, 1.336 (1.323, 1.349) for HF, 1.307 (1.278, 1.337) for VTE, 1.181 (1.163, 1.200) for Afib, and 1.227 (1.218, 1.236) for Death. Figure 2 contains the estimated hazard ratios associated with increasing PM_2.5_ from 5 μg/m^3^ to 10 μg/m^3^ and the estimated PM_2.5_ hazard ratios while receiving corticosteroid therapy.

**Figure 2.**
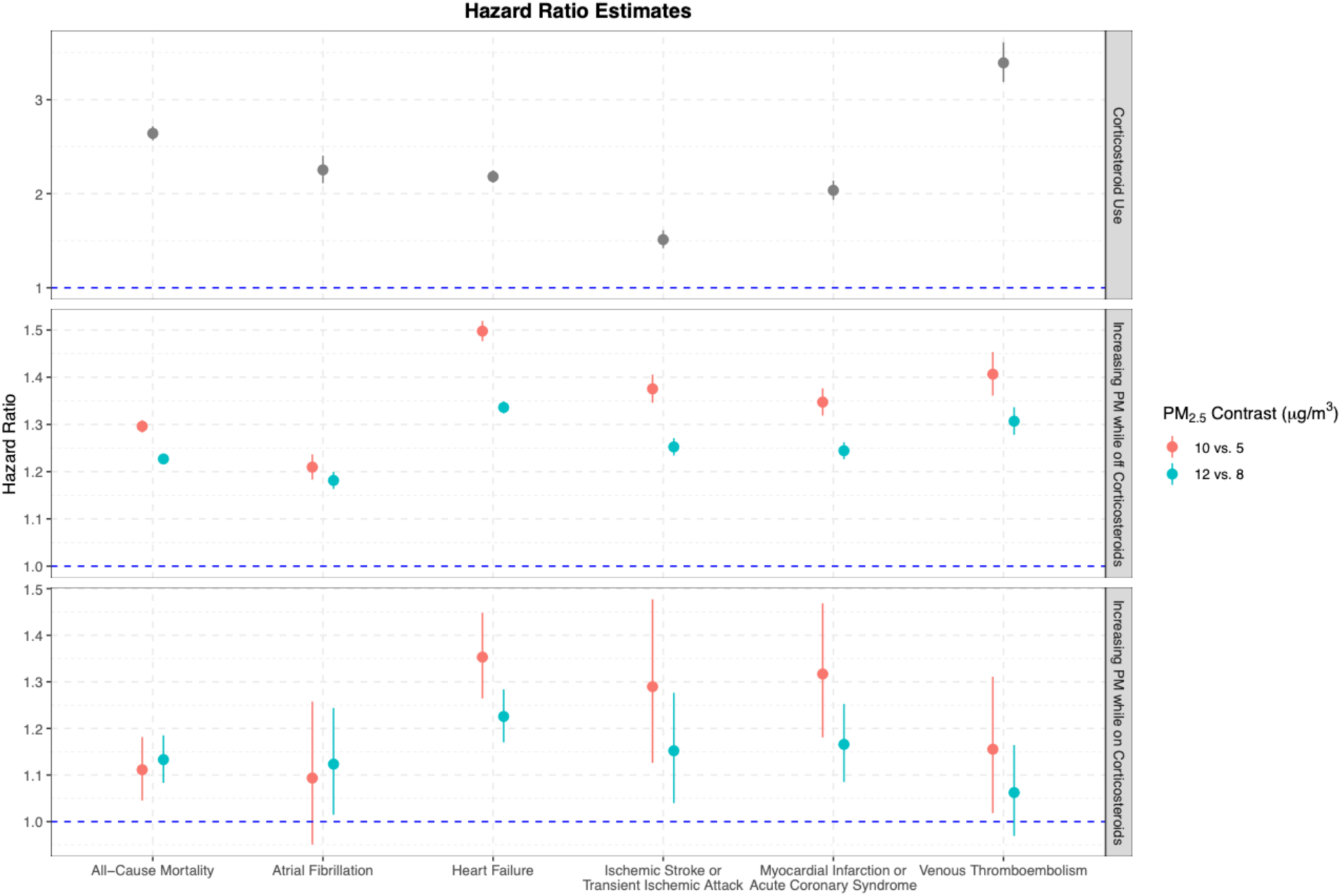
Hazard ratios (and 95% confidence intervals) for corticosteroid use and increasing PM_2.5_ while both on and off corticosteroids. We contrasted the effects of setting PM_2.5_ concentrations to 10 vs. 5 μg/m^3^ and 12 vs. 8 μg/m^3^ which were chosen based on WHO and EPA guidelines.^23,24^

Evaluating the interactions between PM_2.5_ and corticosteroid use on the additive scale, we observed significant interactions (RERI [95% CI]) associated with increasing PM_2.5_ from 8 μg/m^3^ to 12 μg/m^3^ for HF (15.6% [4.0%, 27.3%]), with a borderline significance interaction detected for death (12.5 [-0.6%, 25.5%]). Increasing PM_2.5_ from 5 μg/m^3^ to 10 μg/m^3^ resulted in a significantly increased excess risk due to interaction (RERI [95% CI]) for HF (32.4% [14.9%, 49.9%]) and MI/ACS (29.8% [5.5%, 54.0%]). Figure 3 plots the RERI curves corresponding to various PM_2.5_ contrasts across the range of observed exposure levels for each outcome. For most outcomes, the increase in RERI is steepest when PM_2.5_ is less than 10 μg/m^3^, indicating more intense synergy between PM_2.5_ and corticosteroids even at PM_2.5_ concentrations below current US annual average PM_2.5_ standards.

**Figure 3.**
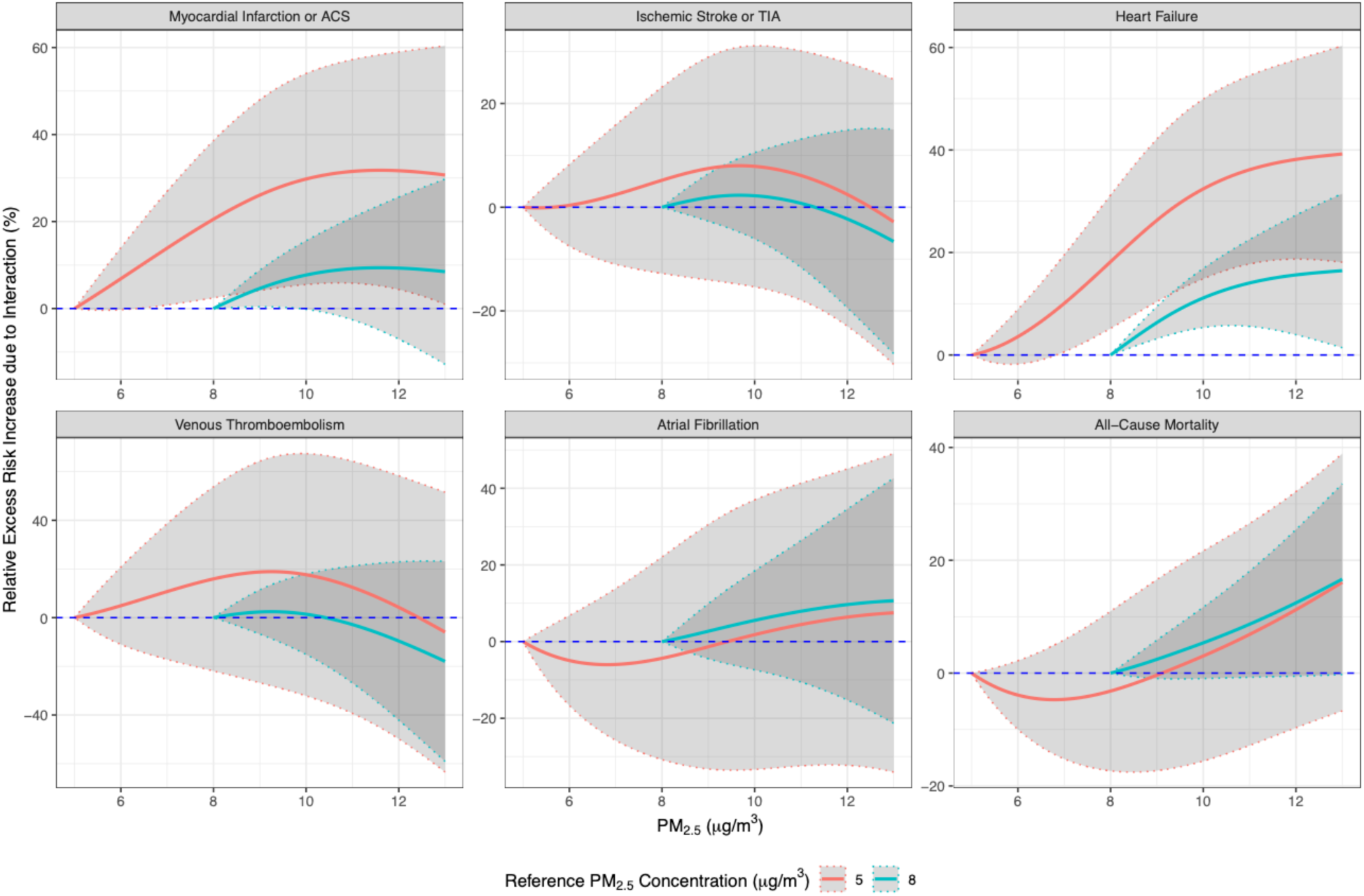
Relative excess risk due to interaction (RERI) between PM_2.5_ and corticosteroid usage for each of the six outcomes (and their 95% confidence interval bands), comparing a range of PM_2.5_ concentrations to reference values of 5 μg/m^3^ (salmon) and 8 μg/m^3^ (blue). Curves represent the change in RERI due to simultaneously initiating corticosteroid treatment and increasing PM_2.5_ exposure to any given level above the corresponding PM_2.5_ reference level. Note that the curves intersect zero at their respective reference levels, as there can be no excess risk increase due to the interaction without changing both exposures concomitantly.

An interesting result worth noting concerns the hazard ratios associated with PM_2.5_ while receiving corticosteroid therapy. Observe that nearly every estimate of the hazard ratio in the third subplot of Figure 2 is attenuated toward the null value of one relative to the second subplot in Figure 2 examining the effects of PM_2.5_ while not receiving corticosteroid therapy. This implies that the multiplicative interaction between PM_2.5_ and corticosteroid use is negative. Thus, our results provide an example of the discordance that can occur between additive and multiplicative measures of synergy, which is well-established in the literature, and further demonstrates the caution one needs to consider when evaluating potential causal interactions.^40^

## Discussion

In this study, we examined the interaction between seasonal-average PM_2.5_ exposure and corticosteroid use on the risk of CTE in a cohort of Medicare beneficiaries with high-risk conditions for CTE from the broadly generalizable set of Fee-for-Service enrollees. Using marginal structural models from the causal inference literature, which adjust for time-varying confounding attributable to several observed neighborhood-and individual-level covariates, we found that the escalation in risk for certain CTE outcomes during periods of simultaneous high PM_2.5_ exposure and corticosteroid use was larger than what would be expected from the independent effects of the two factors added together. In particular, we detected synergism between these two exposures for heart failure and myocardial infarction/ACS.

Numerous studies have reported that older adults and those with comorbidities, particularly respiratory and cardiovascular disease,^41^ are at elevated risk for mortality and morbidity from air pollution. Older adults may be more vulnerable not only because of age and pre-existing diseases, but also because of the multiple medications they receive.^42^ Despite making up only 13% of the US population, older adults account for more than one-third of all prescriptions dispensed.^43,44^ Yet, current evidence on the health impacts of air pollution lacks consideration of additional factors to characterize individuals at risk.^40,45^ In particular, studies lack considerations for medication use, a prevalent risk factor that may further increase vulnerability in older adults.^45^ To our knowledge, our study provides the first epidemiologic evidence of synergistic effects of air pollution and medication on CTE outcomes in older adults.

In addition, examining the independent effects of PM_2.5_ and corticosteroid use on CTE and mortality, we observed results that corroborate those already found in the current literature. PM_2.5_ has been significantly associated with increased risk of MI/ACS,^6^ Stroke/TIA,^7^ HF,^46^ VTE,^14^ Afib,^5^ and all-cause mortality.^47^ Our results sometimes yielded associations larger in magnitude than those found in previous studies. This is unsurprising given that our cohort consists only of participants already at high-risk for CTE.^14^ Likewise, corticosteroid use was strongly and significantly associated with increased risk of the five CTE outcomes and all-cause mortality in our study. The deleterious effects that corticosteroids can have on CTE outcomes have already been described in several other reports.^19,20^

There are several potential biological mechanisms explaining the synergistic interactions between prescription systemic corticosteroids and PM_2.5_ on CTE. First, both PM_2.5_ and glucocorticoids have been shown to induce hypercoagulable states in humans. As PM_2.5_ is small enough to translocate into the bloodstream, chronic PM_2.5_ exposure may increase coagulability indirectly through production of pro-oxidative and pro-inflammatory factors that can then induce production of coagulation factors and fibrinogen. Steroids may complement this thrombogenicity by stimulating Plasminogen Activator Inhibitor-1, which decreases dissolution of fibrinogen.^48^

Second, PM_2.5_ may also lead to atherosclerotic changes and autonomic cardiac dysfunction (i.e., reduced heart rate variability), which in conjunction with adverse metabolic changes seen with systemic glucocorticoid use, can increase risk of cardiovascular disease-related outcomes. Third, both PM_2.5_ inhalation^49^ and glucocorticoids^50^ have been shown to have vasoconstrictive effects, which can increase blood pressure, risk of hypoxia in cardiac/brain tissue, and ultimately lead to MI or stroke. Some steroids also exhibit mineralocorticoid activity at higher doses which can lead to fluid retention and potassium efflux.^51^ In combination with PM_2.5_’s effects on autonomic dysfunction and modulation of vascular tone,^52^ this could potentially exacerbate heart failure or induce arrhythmias.

Our analysis is not without its limitations. First, using model-based PM_2.5_ aggregated to ZIP-codes carries the potential for attenuation created by exposure measurement error.^53^ However, even with such potential attenuation, we still obtained significant results. Second, comorbidities were captured and fixed at the index date and not allowed to vary over time. However, most comorbidities that we accounted for are chronic diseases that are rarely reversed. Third, we censored participants after their first corticosteroid therapy ended, and repeated corticosteroid exposure was not considered in the analyses. A recurrent events model might have been constructed to alleviate this issue; however, fitting marginal structural models in this design is both more time-intensive and new to the causal inference space. Moreover, our approach to consider the first course of exposure makes epidemiological sense given that repeated drug exposures are likely to be associated with worsening of the disease or comorbidities, which is difficult to correct for in a model of the exposure responses.^54^ Fourth, exposure to PM_2.5_ may contribute to the development of diseases that necessitate corticosteroid therapies, such that corticosteroids may mediate the overall impact of PM_2.5_. However, in this paper we did not specifically assess this role of corticosteroids as a potential mediator of PM2.5 effects. Instead, our focus was on examining the combined effects of PM_2.5_ and corticosteroid use. Finally, while we allowed for differing baseline risks of the outcomes for certain disease indications, we did not investigate the potential differential exposure effects experienced by distinct groups of patients using corticosteroids. Based on the findings of this initial analysis, further investigation of these heterogeneous effects is necessary in future studies.

## Conclusion

Using a cohort of nearly two million adults at high-risk for CTE, we found evidence of a synergistic effect between seasonal PM_2.5_ exposure and corticosteroid use on several CTEs.

We used advanced causal inference methods to control for potential confounding attributable to a large set of individual-and neighborhood-level covariates. We also observed strong independent impacts of PM_2.5_ and corticosteroids on each of the six outcomes examined. Our study demonstrates that certain combinations of medication and PM_2.5_ can work synergistically to impose increased health risks on older adults, even when PM_2.5_ concentrations fall below EPA standards. While our results should not discourage clinicians/older adults from prescribing/taking medications needed for treatment, they do shape our conceptual model of disease risk, which we believe should incorporate potential synergisms between individual-and environmental-level risk factors. Our results also emphasize the need for stricter control of PM_2.5_ concentrations to help protect these vulnerable populations for whom corticosteroid medications are commonly indicated.

## Contributors

KPJ, RCN, and SS conducted the analysis, interpreted the data, and drafted the article. AV, BB, AP, and DR were responsible for writing portions of the literature review and aided in drafting the introduction and discussion of the article, particularly the clinical consequences of the results. PG, AP, and MR were responsible for preprocessing and collating the data, along with other data management and administrative duties. SS guided and supervised the research process. RCN and SS provided funding support. All authors provided critical feedback on the study design and revised the manuscript. All authors approved the final draft of the manuscript and are accountable for all aspects of the work reported herein.

## Ethics approval

The study was approved by the Rutgers Institutional Review Board (ID Pro20170001685).

## Data Availability

For data privacy reasons, the Medicare data used in this study cannot be made publicly available, but interested parties can request access by applying through the US Centers for Medicare and Medicaid Services. The PM2.5 exposure data are publicly available at the following link: https://doi.org/10.7927/0rvr-4538. Area-level covariates used herein are also publicly available from the US Census Bureau website. Code for fitting the models and for plotting estimates found within this manuscript is available at: https://github.com/kevjosey/pm-steroid.

