## Supplement for "A retrospective cohort study investigating synergism of air pollution and corticosteroid exposure in promoting cardiovascular and thromboembolic events in older adults"

### Additional Details on the Statistical Models

Let $i = 1,2, \dots, n$ index study participants, $j = 1,2, \dots, m_i$ index individual-level drug-quarters, $k = 1,2, \dots, l$ index ZIP-codes, and $s = 1,2, \dots, 36$ index seasons from Winter 2008 ($s = 1$) to Fall 2016 ($s = 36$). We will denote the $PM_{2.5}$ measurements by $W$ and the corticosteroid indicators by $X$. Define $U$ to be the confounders for $PM_{2.5}$ indexed by $k$ and $s$, and $V$ to be the confounder measurements for corticosteroid therapy indexed by $i$ and $j$. The variable $\underline{W}$ is the vector of four season averaged $PM_{2.5}$ measurements occurring before the present season and the variable $\underline{X}$ denotes the treatment history one quarter prior to the current drug quarter. Lower-case letters refer to the observed values of the corresponding upper-case random variables.

Below we outline the steps to constructing the inverse probability weights used to fit the Cox proportional hazard model which allows us to evaluate the independent and synergistic effects of $PM_{2.5}$ and corticosteroid use.

**Step 1:** Inverse Probability Weighting

Fit the ZIP-code by season-level generalized propensity score for $PM_{2.5}$ with gradient boosting regressions assuming:

$$p_{ks} = \frac{f_W\left(w_{ks} | \underline{W} = \underline{w}_{ks}\right)}{f_W\left(w_{ks} | U = u_{ks}, \underline{W} = \underline{w}_{ks}\right)}$$

Here, $f_W(\cdot)$ denotes the probability density function of the $PM_{2.5}$ measurements evaluated at the observed $PM_{2.5}$ values. Next, we fit an individual- by quarter-level propensity score for corticosteroid use with gradient boosting classification. Taking the cumulative product of the resulting probabilities, we define:

$$q_{ij} = \prod_{\{h:h\leq j, \underline{x}_{ih}=0\}} \frac{Pr\{X = x_{ih} | W = w_{ih}\}}{Pr\{X = x_{ih} | U = u_{ih}, V = v_{ih}, W = w_{ih}\}}.$$

Note that the index on the generalized propensity score weights from $PM_{2.5}$ exposures and the neighborhood-level confounders change to the individual-level indexes in this model. The observations $w_{ij}$ and $u_{ij}$ simply refer to the seasonal average $PM_{2.5}$ and ZIP-code measurements that person $i$ experiences at the start of quarter $j$. Finally, we fit individual by quarter-level probabilities of being censored with gradient boositng (e.g. censoring weights):

$$r_{ij} = \prod_{\{h:h\leq j\}} \frac{Pr\{Censored = 0 | W = w_{ih}, X = x_{ih}\}}{Pr\{Censored = 0 | \, U = u_{ih}, V = v_{ih}, W = w_{ih}, X = x_{ih}\}}.$$

The three inverse probability weights are combined into one weight by finding the product $a_{ijks} = p_{ks} \times q_{ij} \times r_{ij}$. Plugging in estimates for the various probability models (like the output from our gradient boosting regressions) contained in $p_{ks}$, $q_{ij}$, and $r_{ij}$ will yield the estimator $\hat{a}_{ijks}$, which we will use to weight the Cox proportional hazards models.

Since the proportional hazards models are fit on the continuous age-time scale, denoted by $t_i$ for participant $i = 1,2, \dots, n$, and the weights are fit on panels spanning the age-time continuum, we must define the functions $j_i(t)$, $k_i(t)$, and $s_i(t)$ that provide the drug quarter, ZIP-code, and season experienced by person $i$ at age $t$, respectively. These functions allow us to obtain the inverse probability weights at some age $t$ denoted with $\hat{a}_{ij_i(t)k_i(t)s_i(t)}$, which can be used within a proportional hazards model. Additionally, weights are truncated to fall within the 1st and 99th percentiles to prevent some highly leveraged observations from driving estimation and inference.

**Step 2:** Cox Proportional Hazard Model

Define $z_0(w, x)$ and $z_1(w, x)$ to be a basis of nonlinear functions evaluated at a $PM_{2.5}$ measurement of $w$ while off and on medication ($x$), respectively. More concisely, set

$$z_x(W, X) = \begin{cases} ps_x(8), & X \neq x \\ ps_x(W), & X = x \end{cases}$$

where $ps_x(\cdot)$ forms a penalized spline basis with four degrees of freedom. For each CTE outcome, our goal is to model the following hazard function given the two exposures for $PM_{2.5}$ and corticosteroid use evaluated at age $t$, which we denote as

$$\lambda_d(t|W(t) = w, X(t) = x) = \lambda_{d,0}(t) exp\left[x\beta_1 + z_0^T(w,x)\beta_2 + z_1^T(w,x)\beta_3\right].$$

$\lambda_{d,0}(t)$ is an unknown baseline hazard function specific to disease indication $d$, a nuisance parameter that makes the Cox proportional hazard model semiparametric. The hazard models described by the above form are fit by maximizing the partial likelihood while conditioning on the time-varying exposures for $PM_{2.5}$, $W_i(t)$, corticosteroid exposure $X_i(t)$, and given the estimated inverse probability weights fit in Step 1.

**Step 3:** Evaluating the Relative Excess Risk due to Interaction

The existence of a sufficient cause interaction can be tested using the estimated hazard functions to construct the relative excess risk due to interaction. For two contrasting $PM_{2.5}$ values $w_0$ and $w_1$, evidence of synergy can be found by evaluating whether

$$RERI_{w_0,w_1} = \frac{\lambda_d(t|W(t) = w_1, X(t) = 1) - \lambda_d(t|W(t) = w_0, X(t) = 1) - \lambda_d(t|W(t) = w_1, X(t) = 0)}{\lambda_d(t|W(t) = w_0, X(t) = 0)} + 1 > 0.$$

Of course, this value can only be estimated from the fitted Cox model, and thus we must also find some estimator of the standard error to discern the level of uncertainty for our estimates to properly evaluate whether the RERI is greater than zero. To find the standard error, we find bootstrap estimates for $\widehat{\beta}_1$, $\widehat{\beta}_1$, and $\widehat{\beta}_3$ in the Cox model fit. To ease the computational burden of the analysis, we use m-out-of-n bootstrap methods where $m = n/log\ log\ (n)$ and $n$ is the number of unique ZIP codes. As we describe in the main manuscript, the RERI for $PM_{2.5}$ and corticosteroid use is a measure of the interaction on the additive scale, which evaluates whether the combined effect of the two exposures is greater than the individual effects added together.

### Additional Tables and Figures

**Table S1.** ICD-9, ICD-10, and CPT codes for cohort-defining conditions

| Condition | ICD-9 codes | ICD-10 codes |
|---|---|---|
| Total Joint Arthroplasty | 81.54, 81.51<br>CPT Code: 27130, 27447 | 0SRC07Z, 0SRC0J9, 0SRC0JA, 0SRC0JZ, 0SRC0KZ, 0SRD07Z, 0SRD0J9, 0SRD0JA, 0SRD0JZ, 0SRD0KZ, 0SRT07Z, 0SRT0J9, 0SRT0JA, 0SRT0JZ, 0SRT0KZ, 0SRU07Z, 0SRU0J9, 0SRU0JA, 0SRU0JZ, 0SRU0KZ, 0SRV07Z, 0SRV0J9, 0SRV0JA, 0SRV0JZ, 0SRV0KZ, 0SRW07Z, 0SRW0J9, 0SRW0JA, 0SRW0JZ, 0SRW0KZ, 0SR9019, 0SR901A, 0SR901Z, 0SR9029, 0SR902A, 0SR902Z, 0SR9039, 0SR903A, 0SR903Z, 0SR9049, 0SR904A, 0SR904Z, 0SR907Z, 0SR90J9, 0SR90JA, 0SR90JZ, 0SR90KZ, 0SRB019, 0SRB01A, 0SRB01Z, 0SRB029, 0SRB02A, 0SRB02Z, 0SRB039, 0SRB03A, 0SRB03Z, 0SRB049, 0SRB04A, 0SRB04Z, 0SRB07Z, 0SRB0J9, 0SRB0JA, 0SRB0JZ, 0SRB0KZ |
| Myocardial Infarction | 410.xx | I21.01, I21.02, I21.09, I21.11, I21.19, I21.21, I21.29, I21.3, I21.4, I22.0, I22.1, I22.2, I22.8, I22.9 |
| Acute Coronary Syndrome | 411.0, 411.1, 411.81, 411.89 | I24.0, I24.1, I24.8, I24.9 |
| All Stroke | 430, 431, 433.xx, 434.xx, 436 | I60.xx, I61.x, I63.xxx, I65.xx, I66.xx |
| Ischemic Stroke | 433.xx, 434.xx, 436 | I63.xxx, I65.xx, I66.xx |
| Transient Ischemic Attack | 435.x | G45.x |
| Atrial Fibrillation | 427.3, 427.31, 427.32 | I48.0, I48.1, I48.2, I48.3, I48.4, I48.91, I48.92 |

| | | |
|---|---|---|
| Venous Thromboembolism | 451.11, 451.19, 451.81, 451.83, 453.2, 453.4, 453.40, 453.41, 453.42, 453.8, 453.82, 453.83, 453.84, 453.85, 453.86, 453.87, 453.89, 415.1, 415.12, 415.13, 415.19 | I80.1, I80.10, I80.11, I80.12, I80.13, I80.20, I80.201, I80.202, I80.203, I80.209, I80.21, I80.211, I80.212, I80.213, I80.219, I80.22, I80.221, I80.222, I80.223, I80.229, I80.23, I80.231, I80.232, I80.233, I80.239, I80.29, I80.291, I80.292, I80.293, I80.299, I82.210, I82.211, I82.220, I82.221, I82.290, I82.291, I82.220, I82.221, I82.401, I82.402, I82.403, I82.409, I82.411, I82.412, I82.413, I82.419, I82.421, I82.422, I82.423, I82.429, I82.431, I82.432, I82.433, I82.439, I82.441, I82.442, I82.443, I82.449, I82.491, I82.492, I82.493, I82.499, I82.4Y1, I82.4Y2, I82.4Y3, I82.4Y9, I82.4Z1, I82.4Z2, I82.4Z3, I82.4Z9, I82.621, I82.622, I82.623, I82.629, I82.A11, I82.A12, I82.A13, I82.A19, I82.B11, I82.B12, I82.B13, I82.B19, I82.C11, I82.C12, I82.C13, I82.C19, I82.890, I26.01, I26.02, I26.09, I26.90, I26.92, I26.99 |
| Carotid Stenosis | 433.1 | I65.23 |
| Peripheral Vascular Disease | 440.20, 440.21, 440.22, 440.23, 440.24, 440.29, 440.30, 440.31, 440.32, 440.4, 443.9 | I70.2xx-I70.7xx, I70.92, I73.9 |
| Cancer | 140.xx-209.xx, 230.xx-239.xx | C00.xx-C96.xx, D10.xx-D49.xx |
| Heart Failure | 428.xx | I50.xx |

**Table S2.** ICD-9 and ICD-10 codes for CTE outcomes

| Condition | ICD-9 codes | ICD-10 codes |
| --- | --- | --- |
| Myocardial Infarction | 410.xx | I21.01, I21.02, I21.09, I21.11, I21.19, I21.21, I21.29, I21.3, I21.4, I22.0, I22.1, I22.2, I22.8, I22.9 |
| Acute Coronary Syndrome | 411.0, 411.1, 411.81, 411.89 | I24.0, I24.1, I24.8, I24.9 |
| Ischemic Stroke | 433.xx, 434.xx, 436 | I63.xxx, I65.xx, I66.xx |
| Transient Ischemic Attack | 435.x | G45.x |
| Atrial Fibrillation | 427.3, 427.31, 427.32 | I48.0, I48.1, I48.2, I48.3, I48.4, I48.91, I48.92 |
| Venous Thromboembolism | 451.11, 451.19, 451.81, 451.83, 453.2, 453.4, 453.40, 453.41, 453.42, 453.8, 453.82, 453.83, 453.84, 453.85, 453.86, 453.87, 453.89, 415.1, 415.12, 415.13, 415.19 | I80.1, I80.10, I80.11, I80.12, I80.13, I80.20, I80.201, I80.202, I80.203, I80.209, I80.21, I80.211, I80.212, I80.213, I80.219, I80.22, I80.221, I80.222, I80.223, I80.229, I80.23, I80.231, I80.232, I80.233, I80.239, I80.29, I80.291, I80.292, I80.293, I80.299, I82.210, I82.211, I82.220, I82.221, I82.290, I82.291, I82.220, I82.221, I82.401, I82.402, I82.403, I82.409, I82.411, I82.412, I82.413, I82.419, I82.421, I82.422, I82.423, I82.429, I82.431, I82.432, I82.433, I82.439, I82.441, I82.442, I82.443, I82.449, I82.491, I82.492, I82.493, I82.499, I82.4Y1, I82.4Y2, I82.4Y3, I82.4Y9, I82.4Z1, I82.4Z2, I82.4Z3, I82.4Z9, I82.621, I82.622, I82.623, I82.629, I82.A11, I82.A12, I82.A13, I82.A19, I82.B11, I82.B12, I82.B13, I82.B19, I82.C11, I82.C12, I82.C13, I82.C19, I82.890, I26.01, I26.02, I26.09, I26.90, I26.92, I26.99 |
| Heart Failure | 428.xx | I50.xx |

**Table S3.** Comorbidities used for adjustment of the IPWs in addition to the variables listed in Tables 1 and 2.

| Comorbidity | All Participants (N = 1,936,786) | | Participants with Autoimmune Diseases (N = 212,697) | | Participants with COPD or Asthma (N = 510,941) | |
|---|---|---|---|---|---|---|
| | N | % | N | % | N | % |
| Acute Renal Failure | 275,481 | 14.22 | 39,782 | 18.70 | 94,224 | 18.44 |
| Atrial Fibrillation | 689,699 | 35.61 | 76,028 | 35.74 | 207,035 | 40.52 |
| Alcohol Abuse | 52,446 | 2.71 | 5,994 | 2.82 | 20,084 | 3.93 |
| Anaphylaxis | 5,495 | 0.28 | 710 | 0.33 | 1,809 | 0.35 |
| Anemia | 1,010,949 | 52.20 | 128,209 | 60.28 | 287,194 | 56.21 |
| Stable Angina | 169,330 | 8.74 | 19,562 | 9.20 | 55,183 | 10.8 |
| Unstable Angina | 170,790 | 8.82 | 18,159 | 8.54 | 53,844 | 10.54 |
| Anxiety | 286,192 | 14.78 | 32,850 | 15.44 | 97,103 | 19.00 |
| Any Mental Disorders | 876,918 | 45.28 | 91,561 | 43.05 | 280,328 | 54.87 |
| Asthma | 231,379 | 11.95 | 30,868 | 14.51 | 180,997 | 35.42 |
| Bipolar | 27,517 | 1.42 | 2,780 | 1.31 | 10,200 | 2.00 |
| Cancer | 644,054 | 33.25 | 71,404 | 33.57 | 173,131 | 33.88 |
| Cardiac Condition | 242,663 | 12.53 | 28,716 | 13.50 | 70,684 | 13.83 |
| Cardiomyopathy | 226,355 | 11.69 | 28,066 | 13.20 | 78,412 | 15.35 |
| Chronic Renal Diseases | 435,069 | 22.46 | 65,753 | 30.91 | 136,379 | 26.69 |
| Chronic Kidney Disease | 270,902 | 13.99 | 44,790 | 21.06 | 84,505 | 16.54 |
| Congenital Heart Failure | 52,511 | 2.71 | 6,226 | 2.93 | 16,789 | 3.29 |
| COPD | 559,867 | 28.91 | 63,771 | 29.98 | 386,992 | 75.74 |
| Other CVD | 184,735 | 9.54 | 18,262 | 8.59 | 51,449 | 10.07 |
| Dementia | 337,130 | 17.41 | 28,015 | 13.17 | 93,136 | 18.23 |

| | | | | | | |
|---|---|---|---|---|---|---|
| Depression | 419,081 | 21.64 | 47,484 | 22.32 | 136,166 | 26.65 |
| Diabetic Peripheral Circulatory Disorder | 99,344 | 5.13 | 11,448 | 5.38 | 30,192 | 5.91 |
| Diabetic Nephropathy | 88,216 | 4.55 | 12,715 | 5.98 | 26,747 | 5.23 |
| Diabetic Neuropathy | 156,100 | 8.06 | 20,765 | 9.76 | 48,351 | 9.46 |
| Diabetic Retinopathy | 69,979 | 3.61 | 8,023 | 3.77 | 18,263 | 3.57 |
| Severe Diarrhea | 244,970 | 12.65 | 33,331 | 15.67 | 77,903 | 15.25 |
| Dissociative Disorder | 358,436 | 18.51 | 49,419 | 23.23 | 104,411 | 20.44 |
| Drug Abuse | 41,986 | 2.17 | 5,717 | 2.69 | 16,172 | 3.17 |
| Drug Allergy | 20,324 | 1.05 | 2,940 | 1.38 | 6,187 | 1.21 |
| Dysrhythmia | 642,491 | 33.17 | 73,734 | 34.67 | 192,707 | 37.72 |
| Dyskinesia | 1,024 | 0.05 | 94 | 0.04 | 349 | 0.07 |
| Edema | 384,581 | 19.86 | 53,592 | 25.20 | 125,063 | 24.48 |
| Electrolyte Imbalance | 751,968 | 38.83 | 90,116 | 42.37 | 233,035 | 45.61 |
| Epilepsy | 27,621 | 1.43 | 2,746 | 1.29 | 7,812 | 1.53 |
| Food Allergy | 1816 | 0.09 | 254 | 0.12 | 638 | 0.12 |
| Hip Fracture | 84,454 | 4.36 | 8,995 | 4.23 | 24,953 | 4.88 |
| Gastrointestinal Disorders | 166,384 | 8.59 | 19,881 | 9.35 | 50,619 | 9.91 |
| Glaucoma | 17,089 | 0.88 | 2,231 | 1.05 | 4,473 | 0.88 |
| Heart Failure | 656,499 | 33.90 | 76,998 | 36.20 | 238,028 | 46.59 |
| HIV | 2,413 | 0.12 | 246 | 0.12 | 778 | 0.15 |
| Hypertension | 1,716,012 | 88.60 | 194,623 | 91.50 | 463,975 | 90.81 |
| Hyperparathyroidism | 1,618 | 0.08 | 345 | 0.16 | 438 | 0.09 |
| Severe Injury | 25,815 | 1.33 | 2,339 | 1.10 | 6,741 | 1.32 |
| Intestinal Diverticulitis | 250,646 | 12.94 | 32,570 | 15.31 | 75,333 | 14.74 |

| | | | | | | |
|---|---|---|---|---|---|---|
| High Lipid Levels | 1,372,612 | 70.87 | 159,259 | 74.88 | 364,830 | 71.4 |
| Liver Disease | 179,097 | 9.25 | 23,015 | 10.82 | 54,904 | 10.75 |
| Prior Myocardial Infarction | 181392 | 9.37 | 17,939 | 8.43 | 56,652 | 11.09 |
| Neutropenia | 22,030 | 1.14 | 2,143 | 1.01 | 5,994 | 1.17 |
| Obese | 317,494 | 16.39 | 46,263 | 21.75 | 98,747 | 19.33 |
| Obstructive Sleep Apnea | 159,762 | 8.25 | 25,516 | 12.00 | 63,329 | 12.39 |
| Osteoarthrosis | 1,103,702 | 56.99 | 147,794 | 69.49 | 283,493 | 55.48 |
| Other IHD | 920,758 | 47.54 | 104,650 | 49.20 | 287,031 | 56.18 |
| Pancreatitis | 27,250 | 1.41 | 3,400 | 1.60 | 8,474 | 1.66 |
| Prior Pneumonia | 355,215 | 18.34 | 39,001 | 18.34 | 162,715 | 31.85 |
| Psychosis | 127,650 | 6.59 | 11,765 | 5.53 | 39,268 | 7.69 |
| PVD | 467,004 | 24.11 | 52,566 | 24.71 | 153,677 | 30.08 |
| Renal Insufficiency | 335,281 | 17.31 | 50,446 | 23.72 | 108,239 | 21.18 |
| Mental Retardation | 969 | 0.05 | 61 | 0.03 | 275 | 0.05 |
| Prior Self Harm | 2,402 | 0.12 | 301 | 0.14 | 738 | 0.14 |
| Sleep Disorder | 310,840 | 16.05 | 43,634 | 20.51 | 111,357 | 21.79 |
| Smoker | 449,206 | 23.19 | 52,359 | 24.62 | 181,134 | 35.45 |
| Prior Stroke | 482,127 | 24.89 | 49,022 | 23.05 | 136,892 | 26.79 |
| Prior Transient Ischemic Attack | 172,988 | 8.93 | 17,376 | 8.17 | 46,015 | 9.01 |
| Ventricular Arrhythmia | 109,956 | 5.68 | 13,472 | 6.33 | 38,207 | 7.48 |
| Hyperthyroidism | 32,822 | 1.69 | 4,002 | 1.88 | 10,104 | 1.98 |
| Hypothyroidism | 499,245 | 25.78 | 61,472 | 28.90 | 138,905 | 27.19 |

**Table S4.** ICD-9 and ICD-10 codes for Cox Model stratification

| Condition | ICD-9 codes | ICD-10 codes |
|---|---|---|
| COPD | 491.xx, 492.xx, 494.xx, 496.xx | J40.xx, J41.xx, J42.xx, J43.xx, J44.xx, J47.xx |
| Asthma | 493.xx | J45.xx |
| Autoimmune | 714.xx, 274.xx, 696.xx, 7100.xx, 555.xx, 556.xx, 7250.xx, 4465.xx, 340.xx, 135.xx, 2830.xx | M05.xx, M06.xx, M45.xx, M08.xx, M09.xx, M10.xx, M07.xx, M32.xx, K50.xx, K51.xx, M353.xx, M315.xx, M316.xx, G35.xx, D86.xx, D590.xx, D591.xx |

**Table S5.** Incidence Rates of Cardiovascular Thromboembolic Events in Overall Cohort and by Systemic Corticosteroid Use.

| Outcome | All | | No Corticosteroid Use | | Corticosteroid Use | |
|---|---|---|---|---|---|---|
| | Person-Years at Risk | Events (per 1,000 Person-Years) | Person-Years at Risk | Events (per 1,000 Person-Years) | Person-Years at Risk | Events (per 1,000 Person-Years) |
| MI or ACS | 4,481,330 | 93,191 (20.8) | 4,398,446 | 89,451 (20.3) | 82,884 | 3,740 (45.1) |
| Ischemic Stroke or TIA | 4,449,201 | 101,611 (22.8) | 4,366,251 | 98,739 (22.6) | 82,950 | 2,872 (34.6) |
| Heart Failure | 4,282,456 | 244,451 (57.1) | 4,204,664 | 232,883 (55.4) | 77,793 | 11,568 (148.7) |
| Venous Thromboembolism | 4,561,261 | 41,635 (9.1) | 4,477,435 | 38,961 (8.7) | 83,826 | 2,674 (31.9) |
| Atrial Fibrillation | 4,395,518 | 118,754 (27.0) | 4,314,107 | 113,727 (26.4) | 81,411 | 5,027 (61.7) |
| Death | 4,629,432 | 491,445 (106.2) | 4,544,200 | 469,449 (103.31) | 85,232 | 21,996 (258.07) |

**Figure S1.** Example of five participants' survival paths over a one-year period. This plot demonstrates how we construct individual-level quarters over which we model corticosteroid therapy. Participants 1, 2, and 4 never receive corticosteroid therapy and thus their quarters align with seasons. An arrow denotes censoring (as in Participants 1 and 4) while an X denotes when an event occurs (as in Participant 2). Participants 3 (pink) and 5 (green) both received corticosteroids, indicated by the respective dots. Note that the quarters are evenly spaced and shifted according to the starting treatment date. The one exception is within the treatment path of Participant 3 – Quarter 1 is shortened by the start of the study period.

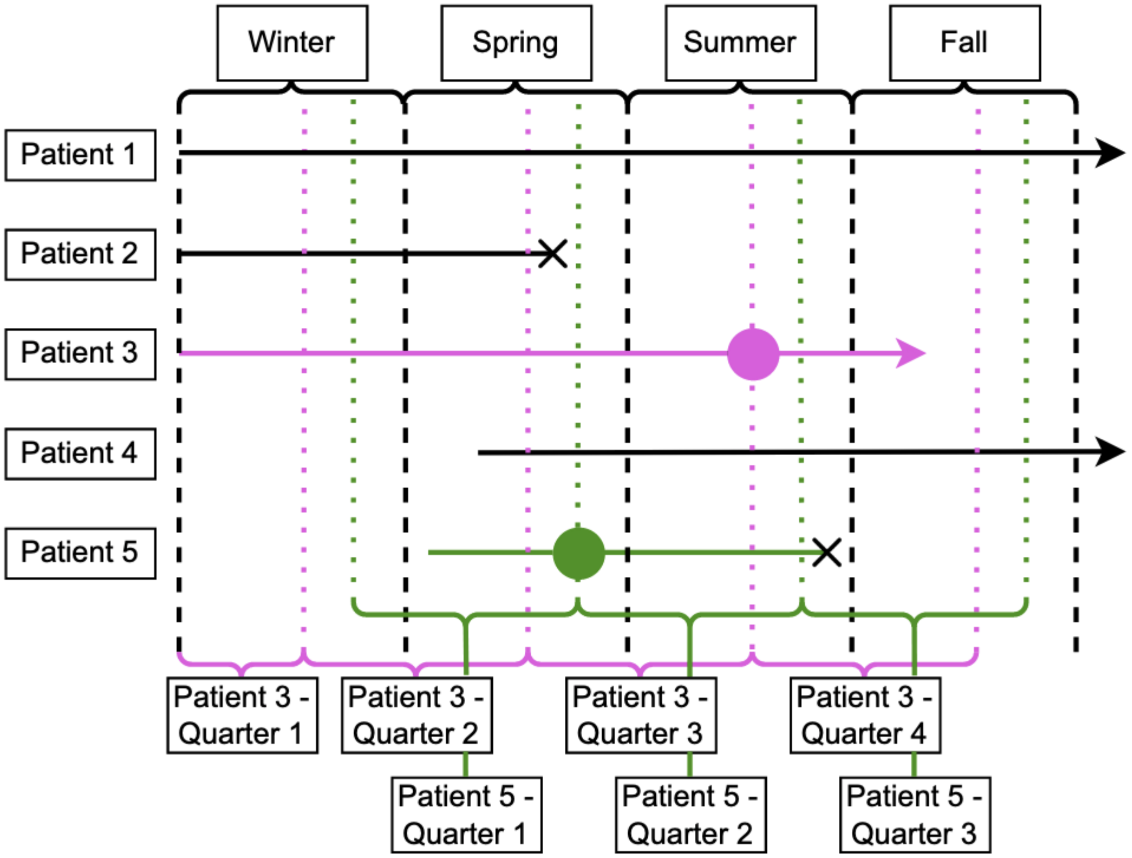
